# Exploring *WNT2* Polymorphisms in Comitant Strabismus: A Genetic Association Study

**DOI:** 10.1101/2024.03.12.24304190

**Authors:** Zainab Zehra, Christopher S. von Bartheld, Andrea B. Agarwal, Hans Vasquez-Gross, Sorath Noorani Siddiqui, Maleeha Azam, Raheel Qamar

## Abstract

**Background:** Strabismus is a complex oculomotor condition characterized by a misalignment of the visual axis. The genetics of strabismus are poorly defined although a few candidate genes have been identified, among which is the WNT2 gene. Our study was designed to assess the association of single nucleotide polymorphisms (SNPs) of *WNT2* in Pakistani strabismus patients.

**Methods:** A total of six SNPs, three intronic and three in the 3’ untranslated region, were screened in the current study. Logistic regression was performed using a dominant, recessive and additive model to determine the association of SNPs with strabismus and its clinical subtypes: esotropia and exotropia. Furthermore, haplotype analysis was performed.

**Results:** Regression analysis revealed an association of rs2896218, rs3779550, rs2285544 and rs4730775 with strabismus under the dominant model. When analyzed separately, rs2896218 and rs2285544 were found to be associated with both esotropia and exotropia, while rs4730775 was significantly associated only with exotropia under the dominant model. Based on clinical parameters, rs2896218, rs2285544 and rs4730775 were also found to be associated with the group of strabismus patients who were diagnosed at birth, but not in the group of patients who were diagnosed later in life. Haplotype analysis revealed that the haplotype A T T (corresponding to rs2896218, rs3779550 and rs2285544) was significantly more prevalent in the strabismus group.

**Conclusion:** Overall, the results of the present study suggests an association of *WNT2* polymorphisms with strabismus and its subtypes in the Pakistani population, though further studies are needed to elucidate their role in strabismus etiology.

**What is already known on this topic**

- Strabismus is a common oculomotor condition with a genetic component.
- *WNT2* has been identified as a candidate gene for comitant strabismus.

**What this study adds**

- Two *WNT2* polymorphisms not previously reported have been found to be associated with strabismus.
- There are genetic variations between clinical subtypes of strabismus (esotropia and exotropia).
- *WNT2* polymorphisms are associated with age at the time of diagnosis and family history.
- Combinations of different alleles (haplotypes) are associated with the disease.

**How this study might affect research, practice or policy**

- Our study adds to the limited genetic data for strabismus and suggests further studies on the role of *WNT2* in strabismus causation.

## Introduction

Strabismus is a common ocular disorder with a worldwide prevalence of 2-3%. The prevalence of horizontal subtypes of strabismus, esotropia and exotropia, vary in different parts of the world and can be influenced by several risk factors such as ethnicity,^1^ seasonality of birth,^2^ gestational age and birth weight, paternal age, and family history.^3^

Comitant strabismus (CS), where the degree of misalignment remains constant regardless of the direction of gaze, accounts for more than 95% of all strabismus cases. Despite being fairly common, the mechanisms underlying its onset are poorly understood. Genetics and a positive family history have been implicated in playing a significant role in the disease. Recent studies have aimed to identify genetic variants responsible for disease causation. In 2018, a genome wide association study (GWAS) by Shabaan et al., (2018)^4^ identified a significant association of rs2244352 in intron 1 of the *WRB* with non-accommodative esotropia in an American population of European ancestry. Two studies identified *LRP2* and *FOXG1* as potential candidate genes for strabismus,^5,6^ while another study suggested *MGST2* and *WNT2* as potential candidate genes for strabismus in a Japanese population.^7^

*WNT2* is a member of the *WNT* family of genes, which encode several secreted signaling proteins. These genes play an important role in early development such as neural development and patterning during embryogenesis,^8^ including early development of ocular tissues.^9^ WNTs are also involved in embryonic muscle development by regulating the activity of myogenic regulatory factors.^10^ WNT2 plays a role in the activation of the canonical WNT signalling pathway by acting as a ligand for Frizzled receptors.^11^ WNT2 can also initiate differentiation of embryonic stem cells via a non-canonical WNT signalling pathway^12^ and may also be involved in the development of embryonic brains.^13^

*WNT2* is located on chromosome 7 (7q31.2), which has previously been identified as a susceptibility locus for CS.^14^ The same group later identified *WNT2* as a candidate gene for CS at this locus. Being an important signalling molecule, together with being a candidate gene for CS, we therefore explored potential roles of the *WNT2* gene by screening its previously reported polymorphisms in a cohort of strabismus patients. The aim of the present study was to investigate whether single nucleotide polymorphisms (SNPs) in *WNT2* are associated with strabismus or its clinical subtypes.

## Methods

### Ethical Review and Cohort Recruitment

Our project was approved by the Ethics Review Board, Department of Biosciences, COMSATS University Islamabad (CUI-Reg/Notif-452/20/526) and conforms to the tenets of the Declaration of Helsinki. All participants and/or accompanying guardians were briefed about the purpose of the study and written consent was obtained prior to sample collection.

Strabismus patients were recruited from the Al-Shifa Trust Eye Hospital, Rawalpindi after diagnosis by an experienced ophthalmologist, which included cover test, prism cover test and Hirschberg test. All participants were briefed about the study before their clinical data were collected. Complete data of all the participants was not available as they or their accompanying guardians had limited information such as, about other strabismus cases in the family or age of the patient at first diagnosis.

### Inclusion/Exclusion Criteria

Patients diagnosed with CS were included in the study regardless of age, gender, or ethnic background. Patients having a history of any other ocular disorder, ocular surgery, trauma or injury to the eye, or any neurological condition were excluded from the study. Patients suffering from phoria, vertical strabismus, paralytic strabismus, and amblyopia-induced strabismus were also excluded. Age-matched healthy controls were recruited from local schools and universities including students and staff.

### Sample Collection and DNA Extraction

A total of 5mL blood was collected from each individual in EDTA tubes and stored at 4°C prior to use. Genomic DNA was isolated from the whole blood using the organic phenol/chloroform method as described by Sambrook and Russel (2001)^15^ with minor modifications.

### Gene and SNP Selection

Previous genetic linkage studies have identified the 7q31.2 gene locus^14^ and *WNT2* gene^7^ as a candidate for strabismus. In our study, we screened a total of six *WNT2* polymorphisms: rs2896218 (NC_000007.14:g.117279924G>A), rs3779550 (NC_000007.14:g.117287306C>T), rs2285544 (NC_000007.14:g.117304229T>A), rs4730775 (NC_000007.14:g.117277064C>T), rs3840660 (NC_000007.14:g.117277191_117277192insA) and rs2024233 (NC_000007.14:g.117277373G>A) in strabismus patients and healthy controls. The SNPs rs2896218, rs2285544 and rs3779550 are intronic variants, while rs4730775, rs3840660 and rs2024233 are present in the 3’ untranslated region (3’ UTR). Three of these variants, rs2896218, rs2285544 and rs2024233 were previously found to be associated with strabismus,^7^ however, the results were not stratified based on clinical subtypes, esotropia and exotropia. Thus, we screened *WNT2* polymorphisms in the Pakistani cohort to ascertain their association with strabismus as well as with its clinical subtypes.

### Genotyping

To screen the selected variants, two methods were employed. Tetra-primer amplification refractory mutation system-polymerase chain reaction (tetra-ARMS-PCR) technique was used for the genotyping of SNPs, rs2896218, rs3779550 and rs2285544. Primers were designed following the protocol as described by Medrano & de Oliveira (2014)^16^ (Supplementary Table 1). About 5% of the samples for each genotype were repeated for cross-verification of the genotyping results. For the other three SNPs, rs4730775, rs3840660 and rs2024233, which were within 500 bp, the Sanger sequencing method was used for genotyping. A total of 66 control and 106 strabismus cases (53 esotropic and 53 exotropic) were sequenced to genotype these three polymorphisms. The sequencing was carried out by the Nevada Genomics Center at the University of Nevada, Reno (https://www.unr.edu/genomics) and the results were viewed using SnapGene Viewer (v.6.2.2). All PCR reactions were done using Q5^®^ Hot Start High-Fidelity 2X Master Mix (New England Biolabs, Inc.). The PCR products were separated on standard agarose gels (2%) in TAE buffer and visualized on a Bio-Rad Gel Doc XR system.

### Statistical Analysis

Hardy Weinberg Equilibrium (HWE) test and Principal Component Analysis (PCA) were performed to test for the population structure of the cohort. Linkage Disequilibrium (LD) was computed to detect the possible linkage between the studied SNPs. Logistic regression analysis under additive, dominant and recessive genetic models was applied where all the SNPs were tested for strabismus (strabismus vs. control), esotropia (esotropia vs. control) and exotropia (exotropia vs. control). In addition, analysis was done after dividing the strabismus group into subgroups based on demographic characteristics including age at diagnosis, family history of strabismus, consanguinity among parents, and exposure to passive smoking. All tests were computed using R statistical software (v.4.2.0).^17^ Haplotypes were generated from the obtained genotypes using Beagle (beagle.08Feb22.fa4.jar). An online tool (https://statpages.info/ctab2x2.html) was used to calculate the odds ratios, 95% confidence interval and p-values between strabismus and controls for each haplotype combination.

## Results

### Cohort overview

A total of 315 strabismic patients (141 esotropic and 162 exotropic) and 230 controls were included in this study. In the strabismus group, 52.9% were males, while in the controls 64.3% were males. The mean age of subjects in the strabismus and control groups was 16.4 and 28.2 years, respectively. Out of the total strabismus cohort 52.1% were diagnosed at birth, 50.9% had a positive family history of strabismus, and 67.1% subjects reported consanguinity among parents (Supplementary Table 2).

### Hardy Weinberg Equilibrium (HWE) and Principal Component Analysis (PCA)

Among the studied variants, only rs4730775, rs3840660 and rs2024233 were found to be in HWE with a p-value of 0.21, 0.76 and 0.54, respectively. However, the genotype frequencies of variants rs2896218, rs3779550 and rs2285544 did not match those expected from the HWE (p-value=< 0.00001). The result from PCA revealed no stratification of the control group in the present cohort with 1000 Genomes data; although, a separate cluster was seen for the strabismus group (Supplementary Figure 1). However, the minor allele frequencies (MAF) of rs2896218, rs3779550 and rs2024233 in the present control group were significantly different from that reported for MAF in the Punjabi cohort from Lahore, Pakistan (PJL) dataset (Supplementary Table 3).

### Linkage Disequilibrium Analysis

Linkage disequilibrium analysis in the control group of the studied cohort revealed linkage between rs2896218, rs3779550 and rs2285544. In addition, the SNPs rs4730775 and rs3840660 were found to be linked, while rs3840660 and rs2024233 were in linkage disequilibrium (Supplementary Table 4).

### Association of *WNT2* polymorphisms with strabismus

Logistic regression analysis under the dominant model showed significant protective association of rs2896218 [Odds Ratio (OR)=0.75 (95% Confidence Interval (CI)=0.66 - 0.86), p-value=5.08e-05] and rs2285544 [OR=0.89 (95% CI=0.81 - 0.97), p-value=0.01], while there was significant risk association of rs3779550 [OR=1.17 (95% CI=1.00 - 1.36), p-value= 0.04] and rs4730775 [OR=1.15 (95% CI=1.00-1.33), p-value=0.05] (Table 1).

**Table 1:** Logistic regression analysis results for the studied variants. Statistically significant p-values are indicated in bold font.

|  |  | Univariable Logistic Regression Analysis |  |  |  |  |  | Logistic Regression Analysis (adjusted for age and gender) |  |  |  |  |  |
| --- | --- | --- | --- | --- | --- | --- | --- | --- | --- | --- | --- | --- | --- |
| SNP | Model | <sup>1</sup>OR<br>(95%CI) | <sup>1</sup>p | <sup>2</sup>OR<br>(95%CI) | <sup>2</sup>p | <sup>3</sup>OR<br>(95%CI) | <sup>3</sup>p | <sup>1</sup>OR<br>(95%CI) | <sup>1</sup>p | <sup>2</sup>OR<br>(95%CI) | <sup>2</sup>p | <sup>3</sup>OR<br>(95%CI) | <sup>3</sup>p |
| rs2896218 | Additive | 0.94<br>(0.87 - 1.02) | 0.14 | 0.88<br>(0.80 - 0.98) | **0.01** | 0.99<br>(0.89 - 1.11) | 0.92 | 0.95<br>(0.89 - 1.03) | 0.21 | 0.92<br>(0.85 - 1.01) | 0.08 | 0.98<br>(0.88 - 1.09) | 0.73 |
|  | Recessive | 1.08<br>(0.96 - 1.20) | 0.21 | 1.07<br>(0.93 - 1.23) | 0.31 | 1.08<br>(0.94 - 1.24) | 0.26 | 1.07<br>(0.97 - 1.19) | 0.18 | 1.09<br>(0.97 - 1.23) | 0.16 | 1.08<br>(0.96 - 1.24) | 0.19 |
|  | Dominant | 0.75<br>(0.66 - 0.86) | **<0.01** | 0.65<br>(0.56 - 0.76) | **<0.01** | 0.81<br>(0.66 - 1.00) | **0.05** | 0.79<br>(0.70 - 0.89) | **<0.01** | 0.71<br>(0.62 - 0.82) | **<0.01** | 0.76<br>(0.62 - 0.92) | **<0.01** |
| rs3779550 | Additive | 1.12<br>(0.97 - 1.28) | 0.12 | 1.09<br>(0.94 - 1.26) | 0.27 | 1.11<br>(0.96 - 1.29) | 0.17 | 1.02<br>(0.91 - 1.16) | 0.72 | 1.00<br>(0.87 - 1.15) | 0.94 | 1.02<br>(0.88 - 1.18) | 0.81 |
|  | Recessive | 0.82<br>(0.55 - 1.23) | 0.35 | 0.89<br>(0.59 - 1.34) | 0.50 | 0.84<br>(0.55 - 1.29) | 0.43 | 0.92<br>(0.63 - 1.33) | 0.64 | 0.94<br>(0.63 - 1.40) | 0.76 | 0.89<br>(0.58 - 1.36) | 0.59 |
|  | Dominant | 1.17<br>(1.00 - 1.36) | **0.04** | 1.12<br>(0.96 - 1.32) | 0.15 | 1.16<br>(0.99 - 1.37) | 0.07 | 1.04<br>(0.91 - 1.19) | 0.58 | 1.01<br>(0.87 - 1.18) | 0.85 | 1.04<br>(0.89 - 1.21) | 0.64 |
| rs2285544 | Additive | 0.92<br>(0.85 - 1.00) | 0.06 | 0.90<br>(0.81 - 0.99) | **0.04** | 0.93<br>(0.85 - 1.03) | 0.17 | 0.94<br>(0.87 - 1.01) | 0.10 | 0.95<br>(0.87 - 1.03) | 0.20 | 0.93<br>(0.85 - 1.02) | 0.12 |
|  | Recessive | 1.31<br>(0.96 - 1.79) | 0.09 | 1.17<br>(0.73 - 1.87) | 0.50 | 1.43<br>(0.99 - 2.05) | 0.05 | 1.10<br>(0.83 - 1.46) | 0.49 | 1.09<br>(0.73 - 1.62) | 0.69 | 1.21<br>(0.87 - 1.68) | 0.26 |
|  | Dominant | 0.89<br>(0.81 - 0.97) | **0.01** | 0.88<br>(0.79 - 0.98) | **0.02** | 0.89<br>(0.81 - 0.99) | **0.04** | 0.92<br>(0.85 - 0.99) | 0.05 | 0.94<br>(0.85 - 1.03) | 0.16 | 0.90<br>(0.82 - 0.99) | **0.04** |
| rs4730775 | Additive | 1.10<br>(1.00 - 1.21) | **0.05** | 1.08<br>(0.95 - 1.22) | 0.22 | 1.14<br>(1.01 - 1.28) | **0.03** | 1.07<br>(0.99 - 1.16) | 0.08 | 1.05<br>(0.95 - 1.16) | 0.34 | 1.14<br>(1.03 - 1.26) | **0.01** |
|  | Recessive | 1.14<br>(0.94 - 1.38) | 0.19 | 1.09<br>(0.84 - 1.41) | 0.52 | 1.22<br>(0.96 - 1.54) | 0.10 | 1.07<br>(0.91 - 1.25) | 0.43 | 1.08<br>(0.88 - 1.33) | 0.48 | 1.17<br>(0.95 - 1.44) | 0.13 |
|  | Dominant | 1.15<br>(1.00 -<br>1.33) | **0.05** | 1.13<br>(0.94 -<br>1.35) | 0.19 | 1.19<br>(1.00 -<br>1.43) | **0.05** | 1.13<br>(1.00 -<br>1.27) | **0.05** | 1.07<br>(0.92 -<br>1.24) | 0.38 | 1.22<br>(1.04 -<br>1.42) | **0.01** |
| rs3840660 | Additive | 1.10<br>(0.99 -<br>1.21) | 0.06 | 1.08<br>(0.95 -<br>1.22) | 0.24 | 1.14<br>(1.00 -<br>1.28) | **0.04** | 1.06<br>(0.98 -<br>1.16) | 0.15 | 1.04<br>(0.94 -<br>1.15) | 0.47 | 1.11<br>(0.99 -<br>1.23) | 0.06 |
|  | Recessive | 1.15<br>(0.95 -<br>1.39) | 0.15 | 1.14<br>(0.89 -<br>1.46) | 0.31 | 1.20<br>(0.95 -<br>1.53) | 0.13 | 1.06<br>(0.90 -<br>1.25) | 0.46 | 1.09<br>(0.89 -<br>1.34) | 0.39 | 1.10<br>(0.89 -<br>1.36) | 0.39 |
|  | Dominant | 1.13<br>(0.98 -<br>1.31) | 0.10 | 1.09<br>(0.91 -<br>1.31) | 0.34 | 1.18<br>(0.99 -<br>1.42) | 0.07 | 1.10<br>(0.97 -<br>1.25) | 0.13 | 1.03<br>(0.89 -<br>1.20) | 0.68 | 1.18<br>(1.01 -<br>1.38) | **0.04** |
| rs2024233 | Additive | 0.97<br>(0.88 -<br>1.07) | 0.56 | 0.97<br>(0.86 -<br>1.09) | 0.66 | 0.97<br>(0.86 -<br>1.09) | 0.58 | 0.97<br>(0.89 -<br>1.04) | 0.42 | 0.94<br>(0.86 -<br>1.04) | 0.30 | 0.97<br>(0.87 -<br>1.07) | 0.54 |
|  | Recessive | 0.98<br>(0.83 -<br>1.15) | 0.78 | 0.96<br>(0.78 -<br>1.18) | 0.70 | 0.99<br>(0.81 -<br>1.21) | 0.93 | 0.97<br>(0.84 -<br>1.11) | 0.62 | 0.93<br>(0.79 -<br>1.10) | 0.41 | 0.99<br>(0.83 -<br>1.19) | 0.94 |
|  | Dominant | 0.95<br>(0.81 -<br>1.11) | 0.50 | 0.97<br>(0.79 -<br>1.18) | 0.74 | 0.92<br>(0.76 -<br>1.12) | 0.41 | 0.95<br>(0.83 -<br>1.08) | 0.40 | 0.93<br>(0.79 -<br>1.09) | 0.37 | 0.92<br>(0.78 -<br>1.09) | 0.35 |

<sup>1</sup> Strabismus vs Controls, <sup>2</sup> Esotropia vs Controls, <sup>3</sup> Exotropia vs Controls
CI, confidence interval; OR, odds ratio

When adjusted for age and gender, rs2896218 [OR=0.79 (95% CI=0.70 - 0.89), p-value= 0.00017], rs2285544 [OR=0.92 (95% CI=0.85 - 0.99), p-value=0.05] and rs4730775 [OR=1.13 (95% CI=1.00 - 1.27), p-value=0.05] retained their significance.

The variant rs4730775 also showed risk association with strabismus under the additive model [OR=1.10 (95% CI=1.00-1.21), p-value=0.05]. However, no association was observed for any of the SNPs under the recessive model.

### Association of *WNT2* polymorphisms with strabismus subtypes

When strabismus subtypes were separately analyzed, a significant protective association was found for rs2896218 and rs2285544 with both esotropia and exotropia under the dominant model. However, rs4730775 showed significant risk association only with exotropia [OR=1.19 (95% CI=1.00 - 1.43), p-value=0.05], which was further strengthened after adjusting for age and gender [OR=1.22 (95% CI=1.04 - 1.42), p-value=0.01].The variant rs3840660 also showed significant risk association with exotropia under dominant model but only after adjusting for age and gender [OR=1.18 (95% CI=1.01 - 1.38), p-value=0.04] (Table 1).

Under the additive model, rs2896218 [OR=0.88 (95% CI)=0.80 - 0.98), p-value= 0.01] and rs2285544 [OR=0.90 (95% CI0.81 - 0.99), p-value= 0.04] showed protective association with esotropia, while rs4730775 [OR=1.14 (95% CI=1.01 - 1.28), p-value= 0.03] and rs3840660 [OR=1.14 (95% CI=1.00 - 1.28), p-value=0.04] were found to be risk associated with exotropia. No association was found between rs2024233 and strabismus or any of its subtypes (Table 1).

### Association of *WNT2* polymorphisms with strabismus based on demographic features

To determine whether the demographic risk factors influenced the association of the studied variants, we stratified the strabismus group based on the age at diagnosis, family history, parental consanguinity and exposure to smoke. The results revealed significant protective association of rs2896218 and rs2285544, and risk association of rs4730775 with strabismus patients diagnosed at birth while no association was observed for any variant with strabismus patients diagnosed later in life (Table 2).

**Table 2:** Univariate regression for strabismus diagnosed at birth and diagnosed later when compared to controls separately (Additive model). Statistically significant p-values are indicated in bold font.

| SNP | Strabismus Diagnosed at Birth vs Control |  |  |  |  |  | Strabismus Diagnosed Later vs Control |  |  |  |  |  |
| --- | --- | --- | --- | --- | --- | --- | --- | --- | --- | --- | --- | --- |
|  | <sup>1</sup>OR<br>(95%CI) | <sup>1</sup>p | <sup>2</sup>OR<br>(95%CI) | <sup>2</sup>p | <sup>3</sup>OR<br>(95%CI) | <sup>3</sup>p | <sup>1</sup>OR<br>(95%CI) | <sup>1</sup>p | <sup>2</sup>OR<br>(95%CI) | <sup>2</sup>p | <sup>3</sup>OR<br>(95%CI) | <sup>3</sup>p |
| rs2896218 | 0.83<br>(0.73 - 0.94) | **<0.01** | 0.84<br>(0.76 - 0.92) | **<0.01** | 0.93<br>(0.82 - 1.05) | 0.23 | 0.97<br>(0.86 - 1.09) | 0.63 | 0.94<br>(0.85 - 1.03) | 0.19 | 1.07<br>(0.97 - 1.17) | 0.21 |
| rs3779550 | 1.13<br>(0.98 - 1.31) | 0.08 | 0.99<br>(0.89 - 1.10) | 0.96 | 1.09<br>(0.96 - 1.26) | 0.19 | 1.07<br>(0.92 - 1.23) | 0.39 | 1.05<br>(0.93 - 1.18) | 0.42 | 1.01<br>(0.91 - 1.14) | 0.80 |
| rs2285544 | 0.89<br>(0.81 - 0.99) | **0.04** | 0.95<br>(0.88 - 1.03) | 0.21 | 0.91<br>(0.83 - 1.00) | 0.06 | 0.93<br>(0.83 - 1.03) | 0.15 | 0.92<br>(0.85 - 1.00) | **0.05** | 1.03<br>(0.95 - 1.11) | 0.51 |
| rs4730775 | 1.14<br>(1.00 - 1.30) | **0.05** | 1.15<br>(1.02 - 1.31) | **0.03** | 1.06<br>(0.93 - 1.21) | 0.38 | 0.97<br>(0.85 - 1.11) | 0.69 | 1.02<br>(0.90 - 1.14) | 0.80 | 0.93<br>(0.84 - 1.04) | 0.22 |
| rs3840660 | 1.13<br>(0.98 - 1.29) | 0.09 | 1.11<br>(0.98 - 1.26) | 0.11 | 1.08<br>(0.94 - 1.23) | 0.28 | 0.99<br>(0.87 - 1.13) | 0.95 | 1.02<br>(0.90 - 1.15) | 0.80 | 0.97<br>(0.87 - 1.08) | 0.59 |
| rs2024233 | 0.96<br>(0.85 - 1.09) | 0.57 | 0.94<br>(0.83 - 1.05) | 0.28 | 1.01<br>(0.89 - 1.14) | 0.94 | 1.09<br>(0.97 - 1.22) | 0.16 | 1.05<br>(0.94 - 1.18) | 0.36 | 1.07<br>(0.97 - 1.17) | 0.17 |
| rs2244352 | 0.98<br>(0.90 - 1.07) | 0.67 | 0.99<br>(0.93 - 1.07) | 0.99 | 0.98<br>(0.91 - 1.07) | 0.69 | 1.03<br>(0.95 - 1.12) | 0.45 | 1.02<br>(0.96 - 1.09) | 0.52 | 1.02<br>(0.96 - 1.09) | 0.46 |

<sup>1</sup> Strabismus vs Controls, <sup>2</sup> Esotropia vs Controls, <sup>3</sup> Exotropia vs Controls
CI, confidence interval; OR, odds ratio

It was also observed that rs2896218 and rs2285544 showed significant protective association, while rs3840660 showed risk association with strabismus patients with a positive family history. The SNP rs3779550 showed protective association with patients with no family history (Table 3). Based on consanguinity, rs2896218 and rs3779550 showed an overall risk association with strabismus from non-consanguineous families. However, rs3840660 showed risk association and rs2024233showed protective association with esotropia in strabismus in consanguineous parents (Table 4). Similarly, no association was observed between the studied variants and strabismus with a positive smoking exposure status. However, rs2285544 showed protective association with overall strabismus with no smoking exposure (Table 5).

**Table 3:** Univariate regression for strabismus cases with positive family history and no family history when compared to controls separately (Additive model). Statistically significant p-values are indicated in bold font.

| SNP | Strabismus with Positive Family History vs Controls |  |  |  |  |  | Strabismus with No Family History vs Controls |  |  |  |  |  |
| --- | --- | --- | --- | --- | --- | --- | --- | --- | --- | --- | --- | --- |
|  | <sup>1</sup>OR<br>(95%CI) | <sup>1</sup>p | <sup>2</sup>OR<br>(95%CI) | <sup>2</sup>p | <sup>3</sup>OR<br>(95%CI) | <sup>3</sup>p | <sup>1</sup>OR<br>(95%CI) | <sup>1</sup>p | <sup>2</sup>OR<br>(95%CI) | <sup>2</sup>p | <sup>3</sup>OR<br>(95%CI) | <sup>3</sup>p |
| rs2896218 | 0.88<br>(0.79 - 0.97) | **0.01** | 0.85<br>(0.77 - 0.94) | **<0.01** | 0.96<br>(0.87 - 1.07) | 0.50 | 0.98<br>(0.88 - 1.09) | 0.68 | 0.92<br>(0.84 - 1.02) | 0.11 | 1.04<br>(0.93 - 1.17) | 0.46 |
| rs3779550 | 1.04<br>(0.90 - 1.19) | 0.60 | 1.00<br>(0.89 - 1.12) | 0.99 | 1.07<br>(0.95 - 1.20) | 0.28 | 1.16<br>(0.99 - 1.36) | **0.05** | 1.09<br>(0.96 - 1.23) | 0.17 | 1.10<br>(0.96 - 1.26) | 0.17 |
| rs2285544 | 0.89<br>(0.82 - 0.99) | **0.03** | 0.89<br>(0.82 - 0.98) | **0.01** | 0.97<br>(0.89 - 1.04) | 0.42 | 0.93<br>(0.84 - 1.03) | 0.15 | 0.95<br>(0.88 - 1.04) | 0.26 | 0.96<br>(0.87 - 1.05) | 0.34 |
| rs4730775 | 1.13<br>(0.99 - 1.29) | 0.06 | 1.06<br>(0.94 - 1.21) | 0.34 | 1.14<br>(1.01 - 1.29) | **0.03** | 1.03<br>(0.90 - 1.17) | 0.71 | 1.06<br>(0.94 - 1.20) | 0.34 | 0.98<br>(0.86 - 1.11) | 0.71 |
| rs3840660 | 1.16<br>(1.02 - 1.32) | **0.02** | 1.09<br>(0.97 - 1.23) | 0.16 | 1.15<br>(1.02 - 1.30) | **0.02** | 0.98<br>(0.86 - 1.13) | 0.80 | 0.98<br>(0.87 - 1.12) | 0.80 | 0.99<br>(0.87 - 1.13) | 0.89 |
| rs2024233 | 0.91<br>(0.81 - 1.03) | 0.14 | 0.92<br>(0.82 - 1.03) | 0.15 | 0.95<br>(0.84 - 1.07) | 0.42 | 1.10<br>(0.97 - 1.25) | 0.12 | 1.10<br>(0.97 - 1.23) | 0.13 | 1.05<br>(0.94 - 1.19) | 0.38 |
| rs2244352 | 0.98<br>(0.90 - 1.07) | 0.68 | 0.99<br>(0.92 - 1.07) | 0.81 | 1.00<br>(0.93 - 1.08) | 0.97 | 1.02<br>(0.94 - 1.11) | 0.60 | 1.02<br>(0.96 - 1.10) | 0.51 | 1.01<br>(0.93 - 1.09) | 0.87 |

<sup>1</sup> Strabismus vs Controls, <sup>2</sup> Esotropia vs Controls, <sup>3</sup> Exotropia vs Controls
CI, confidence interval; OR, odds ratio

**Table 4:**
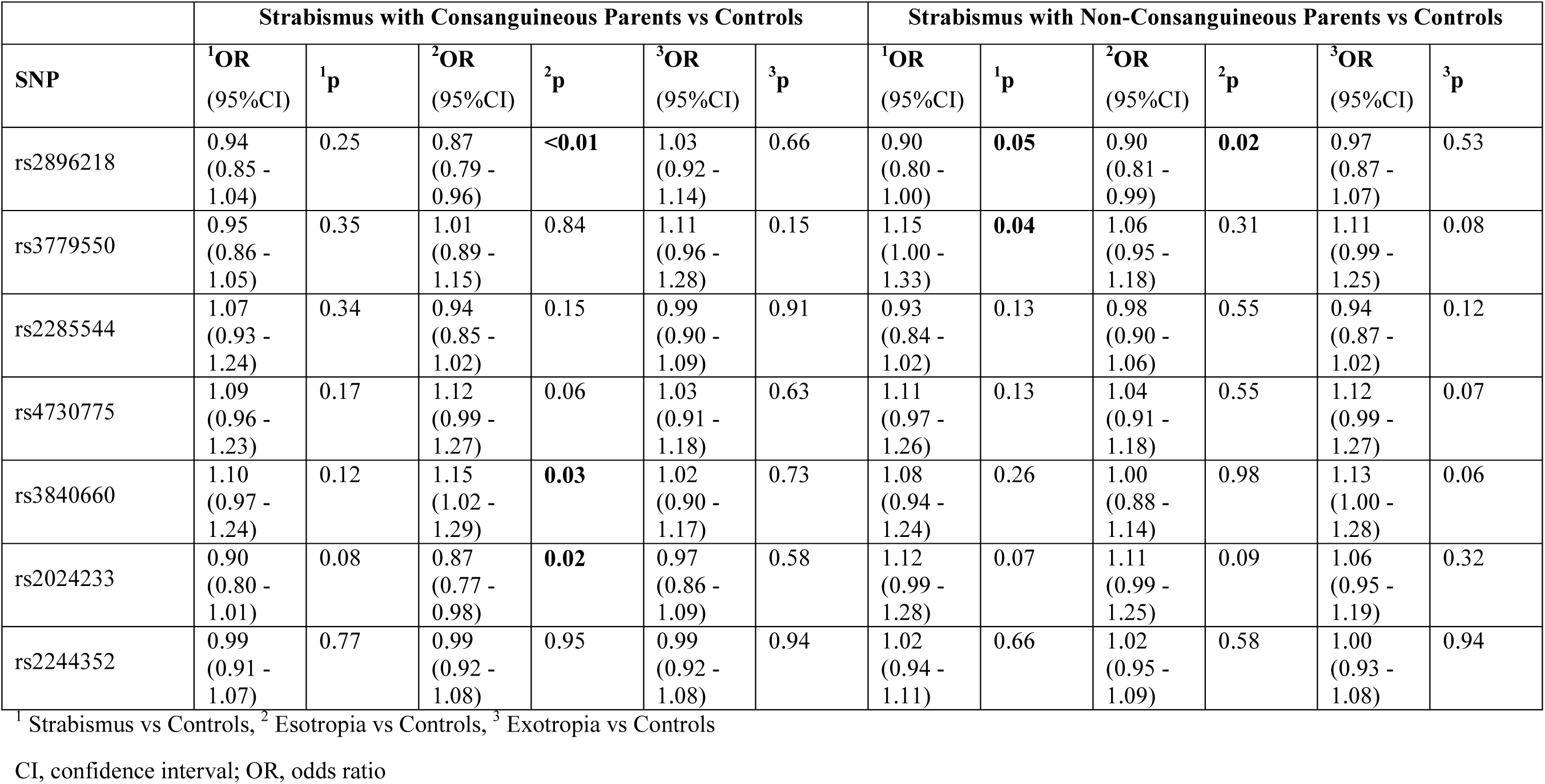
Univariate regression for strabismus cases whose parents had a consanguineous marriage and those with non-consanguineous marriage when compared to controls separately (Additive model). Statistically significant p-values are indicated in bold font.

|  | Strabismus with Consanguineous Parents vs Controls |  |  |  |  |  | Strabismus with Non-Consanguineous Parents vs Controls |  |  |  |  |  |
| --- | --- | --- | --- | --- | --- | --- | --- | --- | --- | --- | --- | --- |
| SNP | <sup>1</sup>OR<br>(95%CI) | <sup>1</sup>p | <sup>2</sup>OR<br>(95%CI) | <sup>2</sup>p | <sup>3</sup>OR<br>(95%CI) | <sup>3</sup>p | <sup>1</sup>OR<br>(95%CI) | <sup>1</sup>p | <sup>2</sup>OR<br>(95%CI) | <sup>2</sup>p | <sup>3</sup>OR<br>(95%CI) | <sup>3</sup>p |
| rs2896218 | 0.94<br>(0.85 -<br>1.04) | 0.25 | 0.87<br>(0.79 -<br>0.96) | **<0.01** | 1.03<br>(0.92 -<br>1.14) | 0.66 | 0.90<br>(0.80 -<br>1.00) | **0.05** | 0.90<br>(0.81 -<br>0.99) | **0.02** | 0.97<br>(0.87 -<br>1.07) | 0.53 |
| rs3779550 | 0.95<br>(0.86 -<br>1.05) | 0.35 | 1.01<br>(0.89 -<br>1.15) | 0.84 | 1.11<br>(0.96 -<br>1.28) | 0.15 | 1.15<br>(1.00 -<br>1.33) | **0.04** | 1.06<br>(0.95 -<br>1.18) | 0.31 | 1.11<br>(0.99 -<br>1.25) | 0.08 |
| rs2285544 | 1.07<br>(0.93 -<br>1.24) | 0.34 | 0.94<br>(0.85 -<br>1.02) | 0.15 | 0.99<br>(0.90 -<br>1.09) | 0.91 | 0.93<br>(0.84 -<br>1.02) | 0.13 | 0.98<br>(0.90 -<br>1.06) | 0.55 | 0.94<br>(0.87 -<br>1.02) | 0.12 |
| rs4730775 | 1.09<br>(0.96 -<br>1.23) | 0.17 | 1.12<br>(0.99 -<br>1.27) | 0.06 | 1.03<br>(0.91 -<br>1.18) | 0.63 | 1.11<br>(0.97 -<br>1.26) | 0.13 | 1.04<br>(0.91 -<br>1.18) | 0.55 | 1.12<br>(0.99 -<br>1.27) | 0.07 |
| rs3840660 | 1.10<br>(0.97 -<br>1.24) | 0.12 | 1.15<br>(1.02 -<br>1.29) | **0.03** | 1.02<br>(0.90 -<br>1.17) | 0.73 | 1.08<br>(0.94 -<br>1.24) | 0.26 | 1.00<br>(0.88 -<br>1.14) | 0.98 | 1.13<br>(1.00 -<br>1.28) | 0.06 |
| rs2024233 | 0.90<br>(0.80 -<br>1.01) | 0.08 | 0.87<br>(0.77 -<br>0.98) | **0.02** | 0.97<br>(0.86 -<br>1.09) | 0.58 | 1.12<br>(0.99 -<br>1.28) | 0.07 | 1.11<br>(0.99 -<br>1.25) | 0.09 | 1.06<br>(0.95 -<br>1.19) | 0.32 |
| rs2244352 | 0.99<br>(0.91 -<br>1.07) | 0.77 | 0.99<br>(0.92 -<br>1.08) | 0.95 | 0.99<br>(0.92 -<br>1.08) | 0.94 | 1.02<br>(0.94 -<br>1.11) | 0.66 | 1.02<br>(0.95 -<br>1.09) | 0.58 | 1.00<br>(0.93 -<br>1.08) | 0.94 |

<sup>1</sup> Strabismus vs Controls, <sup>2</sup> Esotropia vs Controls, <sup>3</sup> Exotropia vs Controls
CI, confidence interval; OR, odds ratio

**Table 5:** Univariate regression for strabismus cases who had intrauterine or early infancy exposure to cigarette smoke and those with no smoking exposure when compared to controls separately (Additive model). Statistically significant p-values are indicated in bold font.

|  | Strabismus with Smoking Exposure vs Controls |  |  |  |  |  | Strabismus with No Smoking Exposure vs Controls |  |  |  |  |  |
| --- | --- | --- | --- | --- | --- | --- | --- | --- | --- | --- | --- | --- |
| SNP | <sup>1</sup>OR<br>(95%CI) | <sup>1</sup>p | <sup>2</sup>OR<br>(95%CI) | <sup>2</sup>p | <sup>3</sup>OR<br>(95%CI) | <sup>3</sup>p | <sup>1</sup>OR<br>(95%CI) | <sup>1</sup>p | <sup>2</sup>OR<br>(95%CI) | <sup>2</sup>p | <sup>3</sup>OR<br>(95%CI) | <sup>3</sup>p |
| rs2896218 | 0.92<br>(0.83 - 1.03) | 0.14 | 0.92<br>(0.85 - 1.00) | 0.06 | 0.97<br>(0.88 - 1.08) | 0.60 | 0.95<br>(0.86 - 1.06) | 0.36 | 0.91<br>(0.82 - 1.00) | 0.06 | 1.02<br>(0.91 - 1.14) | 0.74 |
| rs3779550 | 1.07<br>(0.94 - 1.22) | 0.30 | 1.01<br>(0.92 - 1.11) | 0.83 | 1.07<br>(0.96 - 1.20) | 0.24 | 1.09<br>(0.94 - 1.26) | 0.24 | 1.04<br>(0.92 - 1.17) | 0.55 | 1.11<br>(0.96 - 1.27) | 0.15 |
| rs2285544 | 0.96<br>(0.88- 1.05) | 0.33 | 0.97<br>(0.91 - 1.04) | 0.41 | 0.97<br>(0.90 - 1.05) | 0.42 | 0.90<br>(0.81 - 0.99) | **0.03** | 0.96<br>(0.88 - 1.05) | 0.35 | 0.91<br>(0.83 - 0.99) | **0.05** |
| rs4730775 | 1.12<br>(0.98 - 1.28) | 0.08 | 1.11<br>(0.99 - 1.24) | 0.07 | 1.05<br>(0.93 - 1.19) | 0.42 | 0.99<br>(0.87- 1.13) | 0.90 | 1.01<br>(0.88 - 1.15) | 0.91 | 0.98<br>(0.86 - 1.11) | 0.72 |
| rs3840660 | 1.08<br>(0.95 - 1.24) | 0.25 | 1.05<br>(0.93 - 1.17) | 0.43 | 1.06<br>(0.94 - 1.19) | 0.36 | 1.04<br>(0.91 - 1.18) | 0.61 | 1.03<br>(0.91 - 1.18) | 0.60 | 1.02<br>(0.90 - 1.15) | 0.79 |
| rs2024233 | 1.03<br>(0.92 - 1.17) | 0.59 | 1.02<br>(0.92 - 1.14) | 0.67 | 1.02<br>(0.92 - 1.14) | 0.68 | 1.01<br>(0.89 - 1.14) | 0.92 | 1.00<br>(0.89 - 1.13) | 0.94 | 1.00<br>(0.90 - 1.13) | 0.94 |
| rs2244352 | 1.01<br>(0.93 - 1.09) | 0.85 | 1.02<br>(0.97 - 1.08) | 0.38 | 0.99<br>(0.93 - 1.06) | 0.84 | 1.02<br>(0.94- 1.11) | 0.69 | 1.03<br>(0.96 - 1.11) | 0.41 | 1.00<br>(0.93- 1.08) | 0.99 |

<sup>1</sup> Strabismus vs Controls, <sup>2</sup> Esotropia vs Controls, <sup>3</sup> Exotropia vs Controls
CI, confidence interval; OR, odds ratio

### Haplotype Analysis

Haplotype analysis was conducted from the obtained genotype data for rs2896218, rs3779550 and rs2285544, to see whether any haplotype combination was associated with strabismus. A total of eight haplotype combinations were obtained with one being a rare haplotype (total frequency = 0.007). It was found that the haplotype A T T (corresponding to rs2896218, rs3779550 and rs2285544, respectively), was significantly more prevalent in the strabismus group with a p-value=0.001 (Table 6). Another haplotype, A T A, was more prevalent in the control group (p-value=0.006). In addition to these, two other haplotypes, G C A and A C A tended to be associated with strabismus and control groups, respectively (p-values=0.05).

**Table 6:** Distribution of haplotypes in strabismus and control groups. Statistically significant p-values are indicated in bold font.

| Rs2896218 | Rs3779550 | Rs2285544 | In Total<br>(n=291)<br>number<br>(frequency) | In Controls<br>(n=155)<br>Number<br>(frequency) | In Strabismus<br>(n=136)<br>Number<br>(frequency) | OR (95% CI) | P-value |
| --- | --- | --- | --- | --- | --- | --- | --- |
| G | T | T | 137 (0.24) | 77 (0.25) | 60 (0.22) | 0.86 (0.58 - 1.26) | 0.43 |
| G | C | T | 110 (0.19) | 66 (0.21) | 44 (0.16) | 0.71 (0.47 - 1.09) | 0.14 |
| A | T | T | 86 (0.15) | 32 (0.10) | 54 (0.20) | 2.15 (1.34 - 3.45) | **<0.01** |
| A | C | A | 78 (0.13) | 50 (0.16) | 28 (0.10) | 0.60 (0.36 - 0.98) | **0.05** |
| A | C | T | 71 (0.12) | 32 (0.10) | 39 (0.14) | 1.45 (0.88 - 2.39) | 0.16 |
| G | C | A | 59 (0.10) | 24 (0.08) | 35 (0.13) | 1.76 (1.02 - 3.04) | **0.05** |
| A | T | A | 37 (0.06) | 28 (0.09) | 9 (0.03) | 0.35 (0.16 - 0.74) | **<0.01** |
| G | T | A | 4 (7.00e-03) | 1 (3.00e-03) | 3 (0.01) | 3.45 (0.36 - 33.36) | 0.34 |

CI, confidence interval; OR, odds ratio; n, number of samples analyzed

## Discussion

Comitant strabismus (CS) is a common condition comprising more than 95% of all strabismus cases. Recent studies have identified candidate genes associated with CS, including *LRP2*, *WRB*, *MGST2*, *WNT*, *FOXG1* and *IGF1*.^4–7,18^ The *WNT2* gene is located at the 7q31.2 locus, which has been previously identified as a susceptibility locus for CS including both of its subtypes, esotropia and exotropia.^14,19^ Later, another study revealed association of *WNT2* polymorphisms, rs2896218, rs2285544 and rs2024233, with strabismus in a Japanese cohort.^7^ However, their results were not stratified based on esotropia and exotropia. In the current study we screened six SNPs of *WNT2* in strabismus patients of Pakistani origin, out of which four were found to be associated with strabismus and one of its subtypes.

Our results revealed a significant protective association of rs2896218 and rs2285544 with esotropia, while disease association was observed for rs4730775 and rs3840660 with exotropia. However, rs2024233 was not associated with any strabismus subtype.

The polymorphism rs4730775 has previously been implicated in fibrosis of musculoskeletal tissues in diseases such as Dupuytren’s disease.^20,21^ Fibrosis of extraocular muscles (EOMs) has also been observed in incomitant forms of strabismus.^22^ While the specific role of *WNT2* in fibrosis of the EOM is not well-established, it is known that the WNT signalling pathway is involved in the regulation of tissue fibrosis in general. WNT signalling has also been implicated in the development of fibrotic conditions in various organs, including the lungs, liver, and skin.^23–25^ Dysregulation of WNT signalling can contribute to the activation of fibroblasts, the production of excessive extracellular matrix (ECM) proteins (collagens), and the development of fibrosis.^26^ Moreover, the ECM stiffness impacts the expression of genes involved in the Wnt/β-catenin pathway, thus resulting in a positive feedback loop.^27^ We have previously reported that the expression of ECM-related genes, especially collagens, are altered in strabismic EOMs,^28^ an observation which was also reported by Agarwal et al., (2016)^29^ and Altick et al., (2012).^30^ ECM also regulates the cell differentiation and mineralization of tissues during craniofacial development.^31^ Barreto et al. (2017)^32^ has shown that stiffer substrates can accelerate bone formation and suggested that ECM stiffness could lead to premature bone ossification in craniosynostosis, a condition characterized by the premature fusion of bones of the skull thus restricting brain growth and development. Aberrations in WNT signalling have also been reported in craniosynostosis,^33–35^ while *WNT2* was found to be dysregulated in Apert syndrome, a genetic condition characterized by fusion of the skull, hands, and feet bones.^36^ Strabismus is the most prevalent ocular abnormality in both non-syndromic and syndromic craniosynostosis, including Apert and Crouzon syndromes.^37^ It is possible that *WNT2*-induced differences in facial and orbital anatomy may contribute to an altered frequency of strabismus and its subtypes.

The prevalence of strabismus is also increased in certain neurological conditions including autism spectrum disorder (ASD) compared to the general population.^38^ A recent study has implicated the involvement of *WNT2* polymorphisms with altered cortical thickness in patients with ASD.^39^Altered cortical thickness has also been reported in CS patients.^40^ Thus, it is possible that errors in early development of the brain may cause strabismus, however further studies exploring brain development in CS are needed to establish the role of *WNT2* polymorphisms in strabismus causation.

A total of six polymorphisms were screened in the present study, out of which only three i.e., rs4730775, rs3840660 and rs2024233 were found to be in HWE. The principle of HWE assumes that there is no mutation, no selection, no gene flow, infinite population size and random mating. Though the sample size for this study was appropriate (according to https://www.calculator.net/) it could be one of the limiting factors in achieving HWE in the studied cohort. Moreover, Pakistan has one of the highest rates of consanguineous marriages (https://www.consang.net/) ^41^ and almost half of our samples have positive consanguinity, which could explain the deviation from HWE, which is based on the premise of random mating. Some of the previous studies from our research group also found that the studied variants did not adhere to the HWE,^18,42^ which could indicate inbreeding and population stratification. However, for the present study, the PCA analysis showed no population stratification for the control cohort used. A study by Salanti et al., (2005)^43^ emphasized that the deviations from HWE in genetic association studies are quite common and, in most cases, can be explained by low sample size. The same study cautioned that the lack of power in the study cohorts could also make it harder to detect the extent of inbreeding and thus any deviation from HWE.

To further validate the protective association of rs2896218 and rs2285544, these variants were found to be benign in terms of disease probability (https://regsnps-intron.ccbb.iupui.edu/). However, the variant rs2285544 was predicted to activate a new cryptic acceptor splice site (https://hsf.genomnis.com/home). This, along with the results of haplotype analysis, further strengthens the possibility of a specific combination of variants and interactions among multiple genetic markers influencing the disease phenotype. Future genetic and proteomic studies on the *WNT2* gene may be helpful in exploring this idea further.

We compared allele frequencies of the studied control cohort to the PJL dataset from the 1000 Genomes project and found significant differences for three of the variants i.e., rs2896218, rs3779550 and rs2024233. Hashmi et al., (2022)^44^ and Khan et al., (2020)^42^ from our research group reported similar observations, one reason for this could be that the majority of the samples collected for this and the previous studies were from the twin cities of Rawalpindi and Islamabad. Both these cities comprise people from diverse backgrounds including a high influx of Pashtuns from Khyber Pakhtunkhwa. The PJL dataset consists of samples from Lahore, which is approximately 400km distant from the twin cities and mostly contains Punjabi ethnic group. The differences of the current cohort from the PJL dataset suggest that the population in the twin cities is heterogenous as compared to the PJL reference dataset. Moreover, the variants screened in the present study are in the non-coding region so their corresponding genotypes in the PJL dataset are derived from low coverage whole genome sequencing data, which does not produce high-quality genotypes.^42^ The number of samples in the PJL dataset is also low as compared to our control cohort, thus further underpowering their results.

The LD pattern obtained for the PJL dataset was compared to the LD calculated from the control cohort of the present study, which also indicated some differences. The controls collected for this study were carefully selected to only include healthy individuals without a history of any ocular or systemic condition, but the same criteria could not be verified for the PJL dataset. Thus, the observed differences could be a result of ‘non-random’ sampling of controls for the present study. The results of our PCA analysis show that the control group did not cluster separately from the overall 1000 Genomes data, confirming that the current control group is a reliable representation of an overall unaffected population. However, we did observe a separate cluster for the strabismus group indicating that this is indeed different from the overall 1000 Genomes data.

The results of the present study suggest an involvement of *WNT2* polymorphisms with strabismus. Being an early developmental gene, it supports the idea that strabismus develops as a result of abnormal muscle architecture and signaling defects during embryonic development. The fact that the association of the studied variants (rs2896218, rs2285544 and rs4730775) was only observed in strabismus cases that were diagnosed at birth and not in the patient group diagnosed later in life, strengthens this conjecture. In addition, the results of our haplotype analysis suggest that the presence and combination of multiple variants may influence the severity of the condition. Nevertheless, many individuals develop strabismus later in life, indicating a heterogenous nature of the disease. It is possible that different underlying mechanisms are at play affecting different stages of muscle development and regeneration capacities or alterations in visual or motor pathways,^45^ ultimately leading to a similar phenotype. Thus, there is a need for extensive genetic studies to further investigate how genetic variations influence strabismus. Moreover, familial studies using high throughput techniques and functional studies must be carried out to identify the genetic variants playing a major role in strabismus causation.

## Data Availability

All data produced in the present work are contained in the manuscript

## Acknowledgments

The authors thank all the participants for their contribution in the study.

---

**Supplementary Figure 1:**
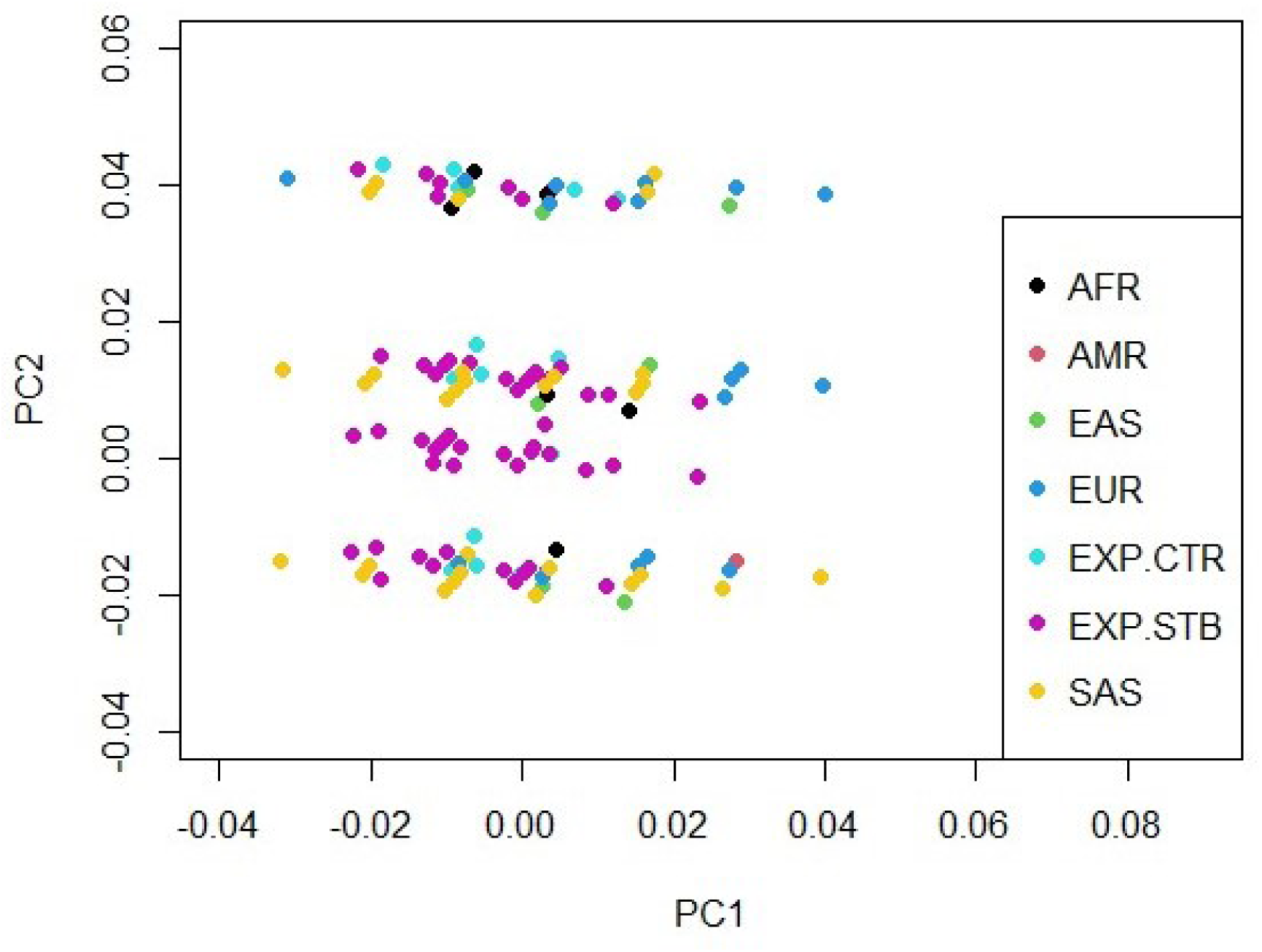
Graphical representation of principal component 1 (PC1) plotted against principal component 2 (PC2) after PCA analysis showing a separate cluster for our strabismus cohort (labeled as EXP.STB), while no stratification is observed for our control cohort (labeled as EXP.CTR) with 1000 Genomes data. AFR, Africans; AMR, Native Americans; EAS, East Asians; EUR, Europeans; EXP.CTR, Experimental Controls; EXP.STB, Experimental Strabismus; SAS, South Asians.

**Supplementary Table 1:**
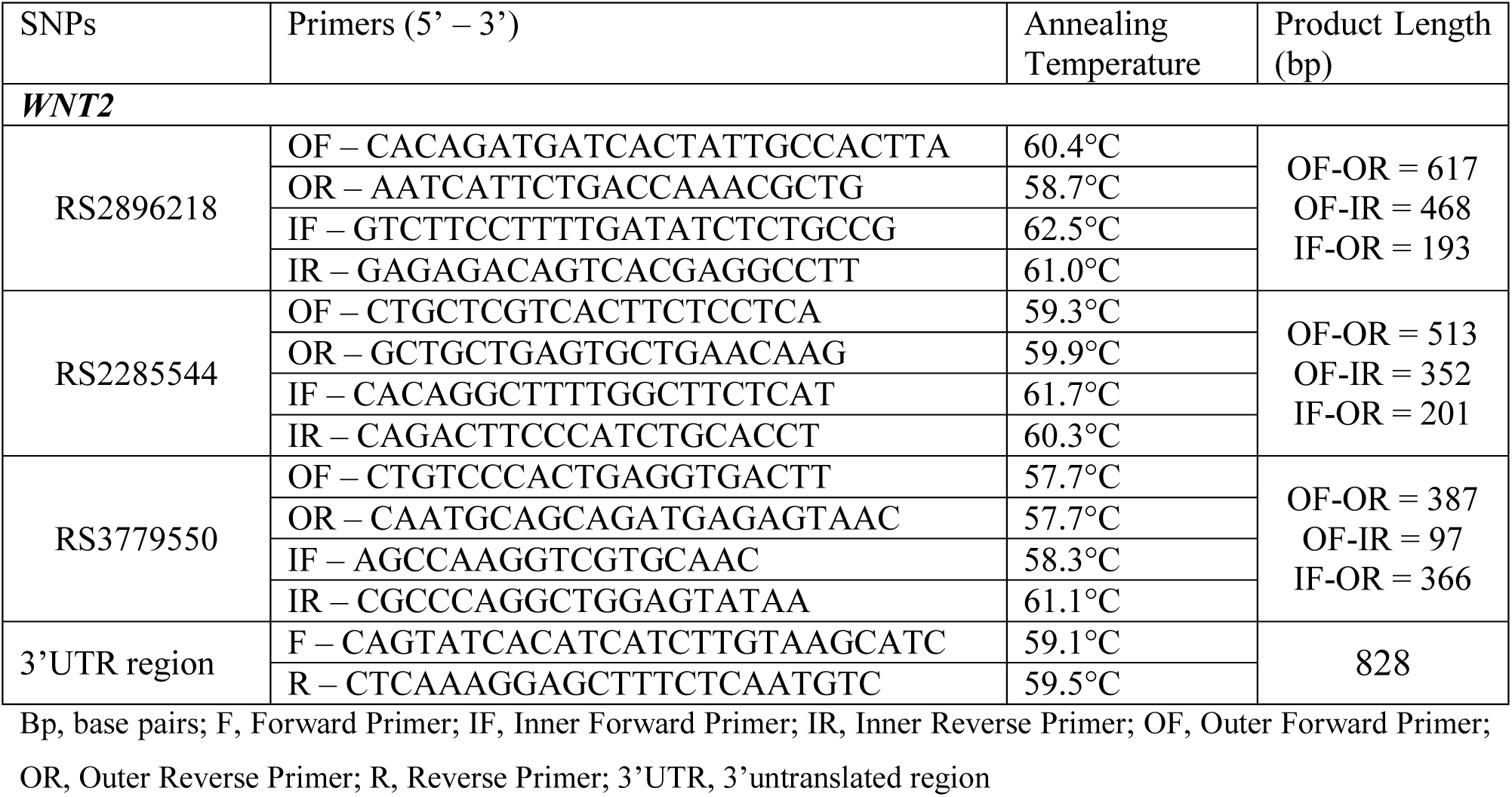
Sequences and details of the primers used in the present study.

**Supplementary Table 2:** Demographic details of the examined cohorts.

| Subjects | Controls | Strabismus | Esotropia | Exotropia |
| --- | --- | --- | --- | --- |
| **Total (N)** | 230 | 315 | 141 | 162 |
| **Gender (Male) (%)** | 64.25 | 52.90 | 52.48 | 53.70 |
| **Age (Years) (mean (±SD))\*** | 28.21 (±12.39) | 16.39 (±9.10) | 14.27 (±8.77) | 18.11 (±9.08) |
| **Diagnosed at Birth**<br>**(Congenital) (%)** | NA | 52.10 | 40.74 | 60.74 |
| **Positive family history (%)** | NA | 50.88 | 55.55 | 47.58 |
| **Parental Consanguinity (%)** | NA | 67.08 | 63.46 | 69.70 |

Values are N for total, percentage (%) for gender and diagnosed at birth, Mean (SD) for age, NA, not applicable.
\*Limitation: Most of the strabismus patients were young children (below the age of 10), but we could not obtain approval of guardians to sample an equal number of unaffected children, thus we used older individuals (ages >14).

**Supplementary Table 3:** Distribution of allele frequencies among the control and disease group along with the calculated and reported minor allele frequencies (MAFs).

| RS Number | Ancestral allele (A1) | Risk allele (A2) | A1 frequency | | A2 frequency | | MAF<sup>1</sup> | MAF<sup>2</sup> | MAF<sup>3</sup> | $\chi^2$ (p-value)<sup>4</sup> | $\chi^2$ (p-value)<sup>5</sup> |
| --- | --- | --- | --- | --- | --- | --- | --- | --- | --- | --- | --- |
|  |  |  | Control (n (%)) | Disease (n (%)) | Control (n (%)) | Disease (n (%)) |  |  |  |  |  |
| Rs2896218 | A | G | 197 (44.97%) | 296 (48.37%) | 241 (55.02%) | 316 (51.63%) | 0.45(A) | 0.48 (A) | 0.62 (A)* | 12.27 (**<0.01**) | 8.32 (**<0.01**) |
| Rs3779550 | C | T | 236 (55.92%) | 248 (53.45%) | 186 (44.07%) | 216 (46.55%) | 0.44 (T) | 0.46 (T) | 0.32 (T) | 6.62 (**0.01**) | 9.01 (**<0.01**) |
| Rs2285544 | T | A | 295 (69.57%) | 361 (74.27%) | 129 (30.42%) | 125 (25.72%) | 0.30 (A) | 0.26 (A) | 0.39 (A) | 3.41 (0.06) | 7.10 (**<0.01**) |
| rs4730775 | C | T | 90 (71.43%) | 127 (59.90%) | 36 (28.57%) | 85 (40.10%) | 0.29 (T) | 0.40 (T) | 0.32 (T) | 0.41 (0.52) | 2.94 (0.09) |
| rs3840660 | No ins | insA | 90 (68.18%) | 122 (57.55%) | 42 (31.82%) | 90 (42.45%) | 0.32 (A) | 0.42 (A) | 0.34 (A) | 0.18 (0.67) | 2.85 (0.09) |
| rs2024233 | A | G | 66 (50.77%) | 114 (54.29%) | 64 (49.23%) | 96 (45.71%) | 0.49 (G) | 0.46 (G) | 0.37 (G) | 6.18 (**0.01**) | 3.48 (0.06) |

<sup>1</sup>MAF calculated from control cohort of present study
<sup>2</sup>MAF calculated from disease cohort
<sup>3</sup>MAF reported for PjL from Ensembl (https://asia.ensembl.org/Homo_sapiens/Info/Index)
<sup>4</sup>Chi-square results for controls and reported data (PjL from Ensembl)
<sup>5</sup>Chi-square results for disease and reported data (PjL from Ensembl)
\* The frequency of A allele was calculated from MAF (G) reported inPjL for rs2896218
Significant p-values (<0.05) are shown in bold font
ins stands for insertion, insA is insertion of A nucleotide
n, number of alleles

**Supplementary Table 4:** Linkage disequilibrium matrix showing D’, R^2^, chi square and p-values for the studied variants for the control cohort. Statistically significant p-values are indicated in bold font.

|  | **rs2896218** | **rs3779550** | **rs2285544** | **rs4730775** | **rs3840660** |
| --- | --- | --- | --- | --- | --- |
| **rs3779550** | D': 0.87<br>R<sup>2</sup>: 0.82<br>Chi-sq: 72.34<br>p-value: **<0.01** |  |  |  |  |
| **rs2285544** | D': 0.85<br>R<sup>2</sup>: 0.76<br>Chi-sq: 61.79<br>p-value: **<0.01** | D': 0.70<br>R<sup>2</sup>: 0.66<br>Chi-sq: 47.26<br>p-value: **<0.01** |  |  |  |
| **rs4730775** | D': 0.19<br>R<sup>2</sup>: 0.13<br>Chi-sq: 1.69<br>p-value: 0.19 | D': 0.11<br>R<sup>2</sup>: 0.08<br>Chi-sq: 0.66<br>p-value: 0.42 | D': 0.08<br>R<sup>2</sup>: 0.06<br>Chi-sq: 0.40<br>p-value: 0.53 |  |  |
| **rs3840660** | D': 0.36<br>R<sup>2</sup>: 0.25<br>Chi-sq: 6.93<br>p-value: **<0.01** | D': 0.08<br>R<sup>2</sup>: 0.06<br>Chi-sq: 0.44<br>p-value: 0.51 | D': 0.07<br>R<sup>2</sup>: 0.05<br>Chi-sq: 0.28<br>p-value: 0.60 | D': 0.85<br>R<sup>2</sup>: 0.79<br>Chi-sq: 68.10<br>p-value: **<0.01** |  |
| **rs2024233** | D': 0.24<br>R<sup>2</sup>: -0.13<br>Chi-sq: 5.02<br>p-value: **0.02** | D': 0.14<br>R<sup>2</sup>: 0.13<br>Chi-sq: 1.73<br>p-value: 0.19 | D': 0.31<br>R<sup>2</sup>: 0.27<br>Chi-sq: 7.85<br>p-value: **<0.01** | D': 0.39<br>R<sup>2</sup>: -0.24<br>Chi-sq: 6.02<br>p-value: **0.01** | D': 0.59<br>R<sup>2</sup>: -0.38<br>Chi-sq: 15.36<br>p-value: **<0.01** |

